# Estimating the direct Disability-Adjusted Life Years (DALYs) associated with SARS-CoV-2 (COVID-19) in the Republic of Ireland: The first full year

**DOI:** 10.1101/2021.12.29.21268120

**Authors:** Declan Moran, Sara Montero Pires, Grant Wyper, Brecht Devleesschauwer, Sarah Cuschieri, Zubair Kabir

## Abstract

**Objectives:** Burden of Disease frameworks facilitate estimation of the health impact of diseases to be translated into a single measure, such as the Disability-Adjusted-Life-Year (DALY).

**Methods:** DALYs were calculated as the sum of Years of Life Lost (YLL) and Years Lived with Disability (YLD) directly associated with COVID-19 in the Republic of Ireland (RoI) from March 01, 2020, to February 28, 2021. Life expectancy is based on the Global Burden of Disease (GBD) Study life tables for 2019.

**Results:** There were 220,273 confirmed cases with a total of 4,500 deaths as a direct result of COVID-19. DALYs were estimated to be 51,532.1 (95% Uncertainty Intervals [UI] 50,671.6, 52,294.3). Overall, YLL contributed to 98.7% of the DALYs. Of total symptomatic cases, 6.5% required hospitalisation and of those hospitalised 10.8% required intensive care unit treatment. COVID-19 was likely to be the second highest cause of death over our study’s duration.

**Conclusion:** Estimating the burden of a disease at national level is useful for comparing its impact with other diseases in the population and across populations. This work sets out to standardise a COVID-19 BoD methodology framework for the RoI and comparable nations in the EU.

## Introduction

By the beginning of March 2020, cases of COVID-19 had been confirmed in most regions throughout the world (1) with the Republic of Ireland (RoI) being no exception. (2) On March 11, 2020, the WHO, declared COVID-19 a pandemic. (3) On March 12, 2020, the Irish government under advice from the National Public Health Emergency Team (NPHET) closed all educational, childcare, and cultural facilities. This initial response was extended to a full lockdown with a stay-at-home order by March 27, 2020. (4) From this date forward Ireland has been living under varying levels of lockdown as the country continues to fight the COVID-19 pandemic. The Department of Health (DoH) in their February 28, 2021, briefing noted that almost all sectors and communities experienced loss and had been “*tested in ways unimaginable to us this time last year”*. (5)

Internationally there has been a wide disparity relating to the direct impact on population health when considering COVID-19. (1) One option to standardise comparisons between countries is to quantify the combined impact of COVID-19’s morbidity and mortality. Burden of Disease (BoD) frameworks facilitate estimation of the health impact of diseases to be translated into a single measure, such as the Disability-Adjusted-Life-Year (DALY). DALYs achieve this through standardising the effects of morbidity and mortality on population health loss as a function of time. The DALY is a health gap metric which measures the healthy life years lost due to diseases, injuries, or risk factors. The DALY is also one of the most internationally used summary measures of population health and is a key metric in the Global Burden of Disease study (GBD). (6) Additionally, the DALY facilitates the comparison of the direct impact of a disease (i.e., COVID-19) against other causes of disease and injury, whilst also facilitating comparison of localised regions or specified demographic groups. (6)

Furthermore, while public conversation on COVID-19 has mainly been centred around those with severe or fatal illness, recent studies highlight an increasing number of people with initially mild COVID-19 who progress to experience prolonged symptoms of which the profile and timeline remains uncertain. Relatively little is known about this “*Long-COVID*”, however, the literature continues to expand. Long-COVID will impact morbidity measures in ongoing and future BoD studies. (7)

Taken together, we set out to estimate the BoD as a direct result of COVID-19 in the RoI from March 01, 2020; to February 28, 2021 (inclusive) using the incidence based internationally comparable metric of DALY.

## Methods

### Data collection

Data relating to COVID-19 in RoI is publicly available through several bodies. Data in this study was sourced from the DATA.GOV.IE website, which collates data from several national organisations, namely the Central Statistics Office (CSO), the Health Protection Surveillance Centre (HPSC), the Health Service Executive (HSE), the Department of Health and the Open Data Unit of the Department of Public Expenditure and Reform (DPER). The data are publicly available.

### Years of Life Lost (YLL)

YLL is the product of the number of deaths (M) and the average remaining life expectancy (RLE) at the time of death: YLL = M x RLE (6)

COVID-19 case definitions in RoI have been routinely updated during the pandemic in accordance with the European Centre for Disease Prevention and Control (ECDC) guidance and updates. (8) Mortality data independently stratified by sex and age-group for the required period was published by the CSO on 10 March 2021 for events created on the Computerised Infectious Disease Reporting (CIDR) system up to midnight Friday 05 March 2021. (9) To exclude data from 01 March 2021 to 05 March 2021 and to allow for mortality reclassification, the DoH daily briefings were reviewed up to and including 31 March 2021. To complete the sex data, the sex ratio from the defined data was applied to the “unknown” cases. To complete the age-group data the age-group ratio from the defined data was applied to the “unknown” cases. Finally, the formulated sex ratio was applied to each age-group.

GBD standard life tables for 2019 were used to calculate remaining life expectancy (RLE) for each sex separately. (10) The average RLE was calculated for each age-group by assuming that the average age of death was the midpoint in each age-group, except for the 80+ category where the average age of death was assumed as the average age of persons in the 80+ category recorded in the Irish Life Tables. (11) Population data was taken from the CSO’s most recent Population and Migration Estimates which were calculated for April 2020. (12)

### Years Lived with Disability (YLD)

YLD was defined as the product of the number of incident cases (N), the average duration until recovery or death (D), and the disability weight (DW), which is reflective of the reduction in one’s health (measured on a scale from 0 (no impact on full health) to 1 (death)): YLDinc = N x D x DW (6)

YLD in this study included 5 health states: asymptomatic, moderate, severe, critical, and post-acute consequences (PAC). Several assumptions were made in relation to informing a person’s categorisation into one of these health states, as seen in Table 1. Polymerase Chain Reaction (PCR) testing criteria for the general public in RoI continues to be based on persons presenting with at least one symptom (being symptomatic), (13) therefore, asymptomatic in this study was defined as a person infected with COVID-19 who did not present for PCR testing; however, this did not account for infections discovered during screening programmes. Asymptomatic cases carried no value for YLD estimation. As described by the European Burden of Disease Network/ECDC consensus disease model, these health states, their description, and their disability weights are based on those from the GBD 2019 study for infectious diseases of the lower respiratory tract, except for those requiring intensive care (Critical), which was defined by the European Disability Weight study. (6, 14) (See table 1)

**Table 1:**
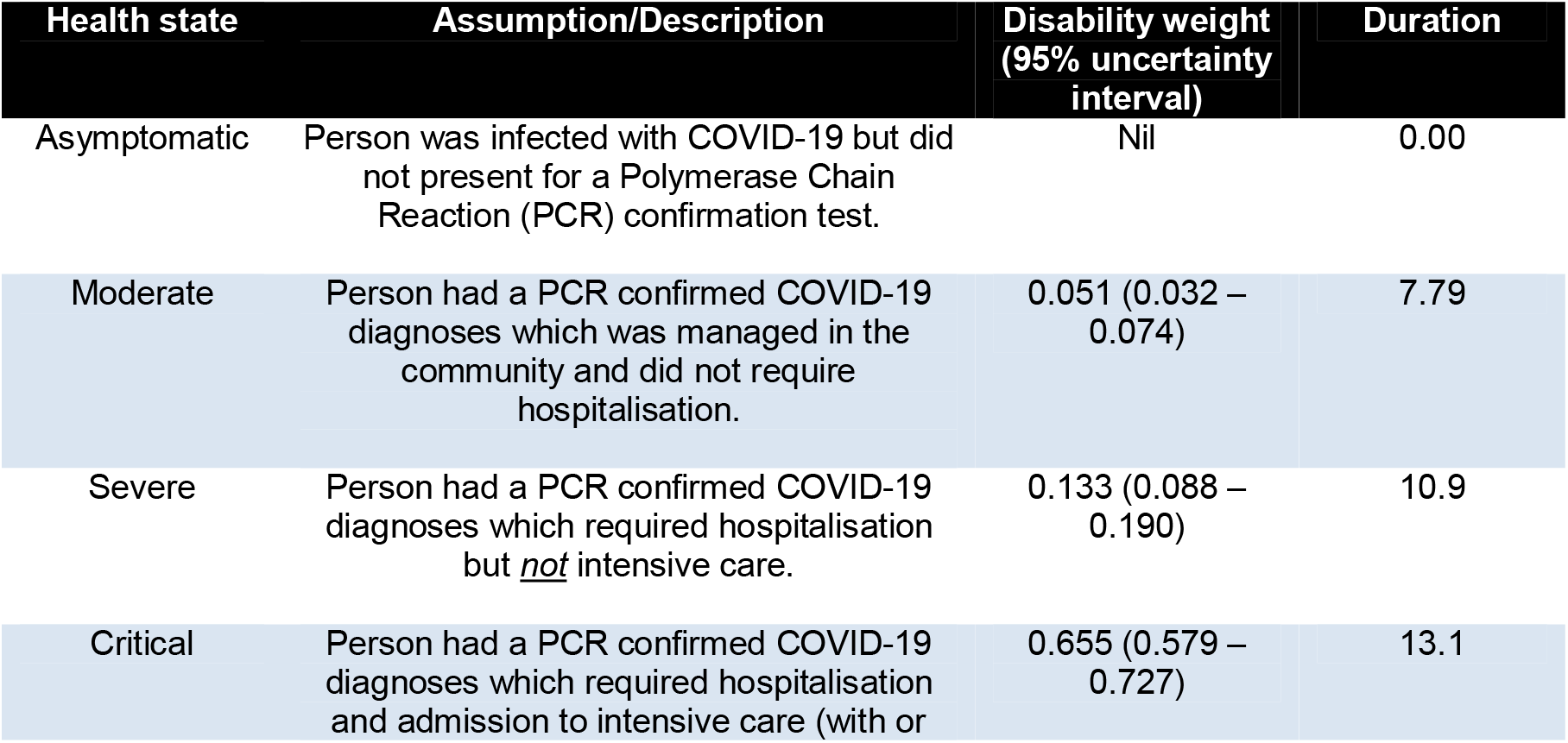
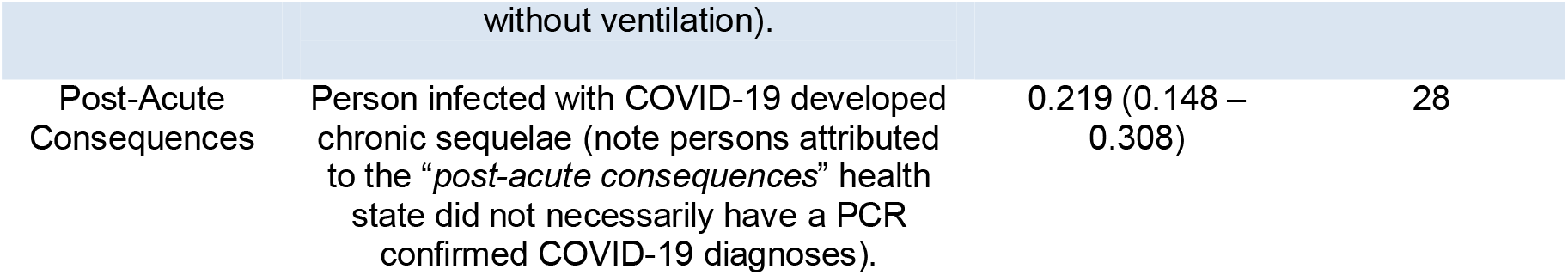
Health state categorisation, disability weights and durations (days). [Burden of disease of COVID-19: Protocol for Country Studies, Brussels, 2020], [Burden of Disease Methods: A Guide to Calculate COVID-19 Disability-Adjusted Life Years. Brussels, 2021]

Morbidity data for all health states, independently stratified by sex and age-group for the required period remains openly available on data.gov.ie (15), except for the health state *“asymptomatic”*, which was assumed to have a 3:1 ratio based on seroprevalence studies. (16, 17) Additionally, the transition probability for the health state “*PAC”* was assumed to be 13.3% of the overall symptomatic incidence with a duration of 28 days. (18) For estimated mean duration relating to health state *“moderate”*, the mean duration for lower respiratory tract infections from the GBD 2019 study was used. (14) Estimated mean duration for health states *“severe”* and *“critical”* were provided by a recent study based on Irish hospital data. (19) Similar to YLL estimates above, a formulated sex ratio was applied to each age-group.

### Disability Adjusted Life Years (DALY)

DALYs are calculated simply by summing the YLLs and YLDs (DALY = YLL + YLD).

### Ethical considerations

Ethical approval for this study was obtained from the Social Research Ethics Committee of University College Cork (UCC).

### Analyses

As DALYs are a unit of time based on person-years, our results are scaled by a factor of 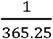, this reflects the contribution of individual days to a complete year. For ease of comparison with previous studies the YLLs, YLDs and DALYs were also estimated per 100,000 population. The uncertainty intervals (UI) from the GBD 2019 life tables were used to estimate the upper and lower intervals of the YLL. The respective disability weight UIs from the GBD 2019 study for infectious diseases of the lower respiratory tract and the European Disability Weight study were used to estimate the upper and lower intervals of the YLD, these UI were subsequently used to estimate the upper and lower estimates of the DALY. Uncertainties were present in our analyses, particularly in relation to the estimation of the proportion of cases who transitioned to health state *“PAC”* and also the duration of health state *“PAC”*. A sensitivity analysis assessed the effect of a 5% and 25% transition probability of symptomatic cases to “*PAC”*. We also assessed *“PAC”* durations of 14 and 56 days. Additionally, a combination of assumptions relating to *“PAC”* were applied to minimise and maximise the YLD. For minimisation we halved the transition probability and the duration of *“PAC”*. For maximisation we doubled the transition probability to *“PAC”*, also applying this to asymptomatic cases and doubled the duration of *“PAC”*. (See table 6).

## Results

### Overall findings

There were 220,273 PCR confirmed cases of COVID-19 with a total of 4,500 deaths (confirmed or probable) as a direct result of COVID-19 within the studies stated parameters. The incidence numbers for each health state are seen in Table 2.

**Table 2:** Incidence by health state.

| Name | Data input proxy | Incidence |
| --- | --- | --- |
| Asymptomatic | Estimate of suspected asymptomatic cases | 660819 |
| Moderate | Confirmed case managed in the community | 205991 |
| Severe | Confirmed case requiring hospital admission (non-ICU) | 12891 |
| Critical | Confirmed case requiring ICU admission | 1391 |
| Post-Acute Consequences | Transition probabilities from cases to post-acute consequences | 29296 |

A fatality rate of 0.49% was estimated when considering all infected persons (asymptomatic + symptomatic). Of symptomatic cases only, we estimate a fatality rate of 2.0% and a hospitalisation rate of 6.5%. Of those hospitalised, 10.8% required treatment in an intensive care unit (ICU).

### Years of Life Lost (YLL)

The sex of each case was defined for 4,154 cases and was “unknown” for 346 cases. Age-group was defined for 4,149 cases and was “unknown” for 351 cases. We estimate YLL of 50,858 (50,220.4, 51,344.8) (males: 24,999.4 (24,652.5, 25,181.3); females: 25,858.6 (25,567.9, 26,163.5)). Based on age, persons in the 65-79 age-groups had the highest YLL at 21,103.9 (20,839.7, 21,368.1) (males: 10,467 (10,326.5, 10,607.5); females: 10,636.9 (10,513.2, 10,760.6)). The highest number of deaths by sex and age-group was seen in males 80+ (1,507, 2143.7 deaths per 100,000 persons). The average age of death was 79 (males: 78.8; females: 79.2). (See table 3).

**Table 3:** Years of life lost by sex and age-group.

| Males | Deaths | YLL | YLL (100,000) |
| --- | --- | --- | --- |
| 0-14 | 0 | 0 | 0 |
| 15-24 | 0 | 0 | 0 |
| 25-44 | 22 | 1,014.9<br>(1,008.3, 1,023.7) | 148 |
| 45-64 | 163 | 4469.3<br>(4420.3, 4501.9) | 735.5 |
| 65-79 | 702 | 10467<br>(10326.5, 10607.5) | 3,895.6 |
| 80+ | 1507 | 9048.2<br>(8897.4, 9048.2) | 12,862 |
| Total | 2394 | 24,999.4<br>(24,652.5, 25,181.3) | 1,013.2 |

| Females | Deaths | YLL | YLL<br>(100,000) |
| --- | --- | --- | --- |
| 0-14 | 0 | 0 | 0 |
| 15-24 | 0 | 0 | 0 |
| 25-44 | 19 | 939.2<br>(933.5, 944.9) | 132 |
| 45-64 | 143 | 4,994.1<br>(4,965.5, 5,037) | 806.3 |
| 65-79 | 618 | 10,636.9<br>(10,513.2, 10,760.6) | 3,785.5 |
| 80+ | 1326 | 9,288.4<br>(9155.7, 9,421) | 9,226.6 |
| Total | 2106 | 25,858.6<br>(25,567.9, 26,163.5) | 1,028.7 |

### Years Lived with Disability (YLD)

Sex was defined in 219,507 cases and was “unknown” in 766 cases. Age-group was defined for 219,466 cases and was “unknown” in 807 cases. We estimate YLD of 674.2 (95% UI 451.1, 949.5). Males constituted YLD of 316.9 (212, 446.3) and females 357.4 (239.1, 503.2). The largest contributing age-group was “*25-44”* with YLD of 229.2 (153.4, 322.8) and the largest contributing health state is “*PAC*” with YLD of 369.1 (249.5, 519.2). The largest contributing combined sub-population (sex, age-group, and health state) were *“females” “25-44” “PAC*” with YLD of 66.5 (45, 93.6), this population consisted of 5,279 persons. (See table 4).

**Table 4:** Years lived with disease estimates.

| ***YLD by health state*** |  |
| --- | --- |
| Health State | YLD |
| Asymptomatic | 0 |
| Moderate | 220.8 (138.5, 320.3) |
| Severe | 51.5 (34.1, 73.5) |
| Critical | 32.8 (29.0, 36.5) |
| Post-Acute Cons. | 369.1 (249.5, 519.2) |
| Total | 674.2 (451.1, 949.5) |

| **YLD by sex and health state** |  |  |  |
| --- | --- | --- | --- |
|  | Health State | Incidence | YLD |
| Males | Moderate | 96,816 | 103.8 (65.1, 150.6) |
|  | Severe | 6,059 | 24.2 (16.0, 34.6) |
|  | Critical | 654 | 15.4 (13.6, 17.1) |
|  | PAC | 10,353 | 173.5 (117.3, 244) |
| Females | Moderate | 109,177 | 117 (73.4, 169.8) |
|  | Severe | 6,832 | 27.3 (18.1, 39) |
|  | Critical | 737 | 17.4 (15.4, 19.3) |
|  | PAC | 11,674 | 195.6 (132.2, 275.2) |
| **YLD by sex and age-group** |  |  |  |
|  | Age-Group | Incidence | YLD |
| **Males** | 0-14 | 10,249 | 28.5 (19.1, 40.2) |
|  | 15-24 | 20,499 | 57 (38.2, 80.3) |
|  | 25-44 | 38,720 | 107.7 (72.1, 151.7) |
|  | 45-64 | 29,609 | 82.4 (55.1, 116) |
|  | 65-79 | 9,110 | 25.4 (17, 35.7) |
|  | 80+ | 5,694 | 15.8 (10.6, 22.3) |
| **Females** | 0-14 | 11,558 | 32.1 (21.5, 45.3) |
|  | 15-24 | 23,115 | 64.3 (43, 90.6) |
|  | 25-44 | 43,663 | 121.5 (81.3, 171.1) |
|  | 45-64 | 33,389 | 92.9 (62.2, 130.8) |
|  | 65-79 | 10,274 | 28.6 (19.1, 40.3) |
|  | 80+ | 6,421 | 17.9 (12, 25.2) |

### Disability Adjusted Life Years (DALYs)

We estimated that COVID-19 caused 51,532.1 DALYs (50,671.6, 52,294.3) in the full year period. Overall, YLL contributed 98.7% towards the DALYs with the remaining 1.3% attributed to YLD. The sub-population with highest DALYs were “*Females” “65-79”* with DALYs of 10,665.5 (10,532.3, 10,800.9), which translates to 3,798.3 (3,750.8, 3,846.5) DALYs per 100,000 persons. However, the largest DALYs per 100,000 persons was seen in the Male 80+ population with a total of 12,893.3 (12,671.4, 12,902.6). We estimated 11.5 (11.3, 11.6) DALYs per death. (See table 5), (See table 6).

**Table 5:** Disability adjusted life years by sex and age-group.

| Age | Males |  | Females |  | Total Persons<br>DALYs |
| --- | --- | --- | --- | --- | --- |
|  | DALYs | DALYs<br>(per100,000) | DALYs | DALYs<br>(per100,000) |  |
| 0-14 | 28.5<br>(19.1, 40.2) | 5.5<br>(3.7, 7.8) | 32.1<br>(21.5, 45.3) | 6.6<br>(4.4, 9.3) | 60.6<br>(40.6, 85.5) |
| 15-24 | 57<br>(38.2, 80.3) | 17.8<br>(11.9, 25) | 64.3<br>(43, 90.6) | 20.7<br>(13.8, 29.2) | 121.3<br>(81.2, 170.9) |
| 25-44 | 1,122.6<br>(1,080.4, 1,175.4) | 163.9<br>(157.7, 171.6) | 1,060.7<br>(1,014.8, 1,116) | 149.1<br>(142.7, 156.9) | 2,183.3<br>(2,095.2, 2,291.4) |
| 45-64 | 4,551.7<br>(4,475.4, 4,617.9) | 749.6<br>(737.1, 760.5) | 5,087<br>(5,027.7, 5,167.8) | 821.8<br>(812.2, 834.9) | 9,638.7<br>(9,503.1, 9,785.7) |
| 65-79 | 10,492.4<br>(10,343.5, 10,643.2) | 3,907.8<br>(3,852.3, 3,963.9) | 10,665.5<br>(10,532.3, 10,800.9) | 3,798.3<br>(3,750.8, 3,846.5) | 21,157.9<br>(20,875.8, 21,444.1) |
| 80+ | 9,064<br>(8,908, 9,070.5) | 9,064<br>(8,908, 9070.5) | 9,306.3<br>(9,167.7, 9,446.2) | 9,250.8<br>(9,113, 9,389.9) | 18,370.3<br>(18,075.7, 18,516.7) |

**Table 6:**
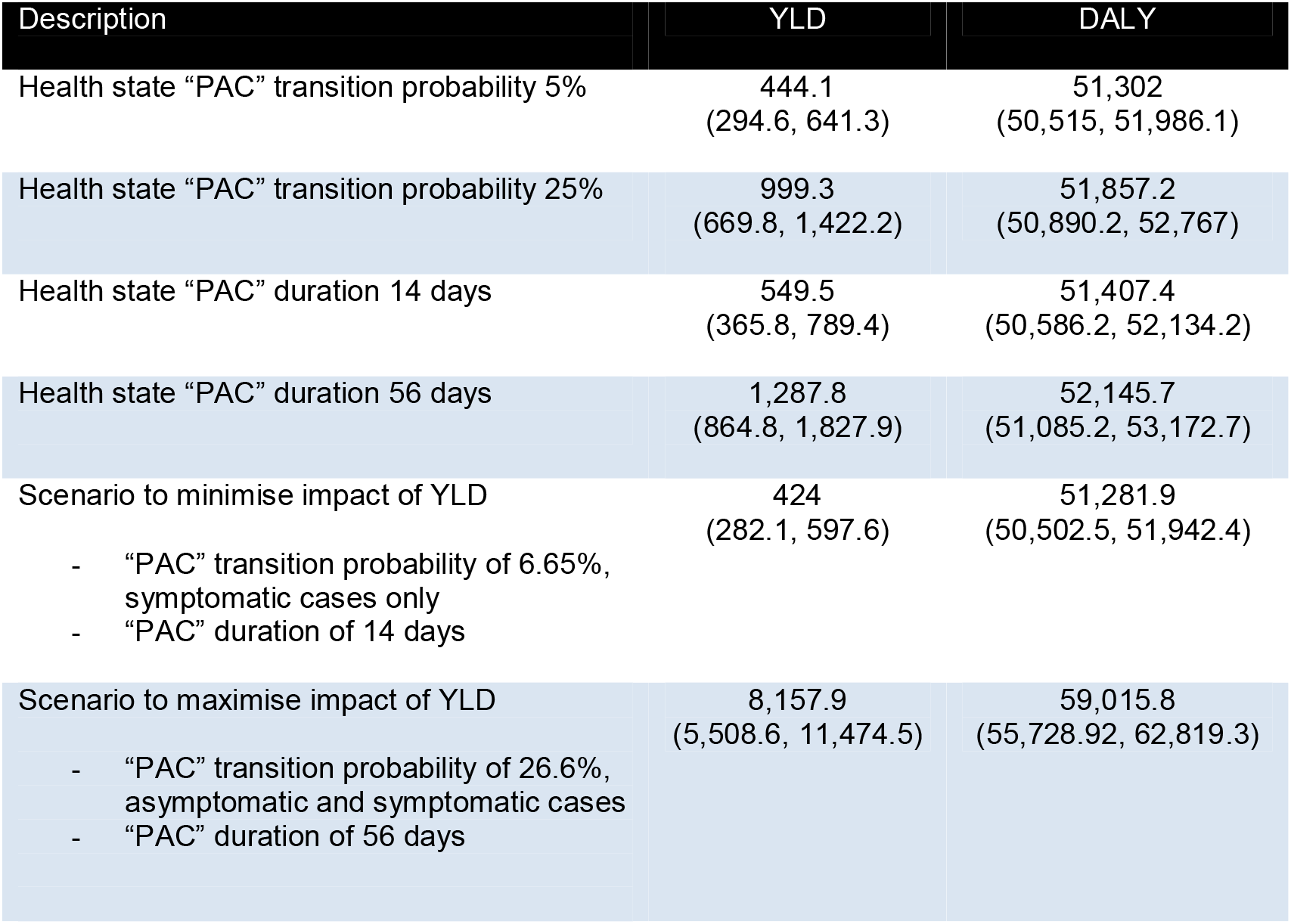
Results of sensitivity & scenario analysis.

| Description | YLD | DALY |
| --- | --- | --- |
| Health state "PAC" transition probability 5% | 444.1<br>(294.6, 641.3) | 51,302<br>(50,515, 51,986.1) |
| Health state "PAC" transition probability 25% | 999.3<br>(669.8, 1,422.2) | 51,857.2<br>(50,890.2, 52,767) |
| Health state "PAC" duration 14 days | 549.5<br>(365.8, 789.4) | 51,407.4<br>(50,586.2, 52,134.2) |
| Health state "PAC" duration 56 days | 1,287.8<br>(864.8, 1,827.9) | 52,145.7<br>(51,085.2, 53,172.7) |
| Scenario to minimise impact of YLD | 424<br>(282.1, 597.6) | 51,281.9<br>(50,502.5, 51,942.4) |
| - "PAC" transition probability of 6.65%,<br>symptomatic cases only |  |  |
| - "PAC" duration of 14 days |  |  |
| Scenario to maximise impact of YLD | 8,157.9<br>(5,508.6, 11,474.5) | 59,015.8<br>(55,728.92, 62,819.3) |
| - "PAC" transition probability of 26.6%,<br>asymptomatic and symptomatic cases |  |  |
| - "PAC" duration of 56 days |  |  |

## Discussion

This study estimated the BoD as a direct result of COVID-19 in the RoI from March 01, 2020; to February 28, 2021 (inclusive) using the incidence-based internationally comparable metric of the DALY. Overall, there were 220,273 PCR confirmed cases with 4,500 deaths (confirmed or probable), and 51,532.1 (95%UI 50671.6, 52294.3) DALYs. The largest impact on population health was a result of premature mortality with YLL representing 98.7% of the DALYs. We estimated 11.3 YLL per death. This is similar to findings in South Korea and the United States with YLL per death of 9.46 and 9.23 respectively, (20, 21) however, other national BoD studies indicated YLL per death ranging from 4.79 to 15.79. (22, 23) Variation in YLL per death across studies is influenced by which life valuation table is applied. Despite extensive population health interventions designed specifically to reduce the risk of COVID-19, deaths directly related to the virus are likely to rank second when compared to RoI 2019 mortality data. The overall DALYs were marginally higher in males than females; this result was also found in a recent United States study. (21) The largest contributing sub-population were females aged 65-79, however, while the largest DALYs per 100,000 persons were in the male 80+ population. DALY contribution significantly increased in populations 65+. This biological inequality suggests that the higher age-groups of both male and female are at a higher risk, particularly of mortality from COVID-19. The results from sensitivity and scenario analyses highlighted that variation in relation to assumptions relating to YLD input variables have minor impact on the overall DALY estimates as YLD contribute a small percentage of the total DALYs.

The DALY metric is a useful health gap summary measure when attempting to identify the impact of a disease on public health. This study adopted the incidence approach for estimating DALYs as incidence data was widely reported.

As the natural history of COVID-19 is evolving, at the time of completion of this study there is no longitudinal data available. DALY estimates for COVID-19 burden are available to a limited number of countries using data over varying time durations. Early on in the pandemic, both South Korea estimated DALYs based on approximately 3-month durations, 2,531 and 121,449 DALYs respectively. (20, 22) Studies of approximately 1 year duration completed in Germany, Scotland, and Malta estimate DALYs of 305,641, 96,519 and 5,478 respectively. (24, 25, 23)

### Contextualisation

For contextualisation, we compared this study’s findings with RoI GBD data for 2019. (26) COVID-19 is likely to be the second highest cause of death in the RoI over our study’s duration, with only ischemic heart disease (IHD) causing more deaths. Due to mortality being such a high contributing factor to the DALYs, COVID-19 is also likely to have the second highest YLL. Additionally, our COVID-19 DALY estimates are comparable in magnitude to estimates due to ‘Unintentional Injuries’ (54,835.6) and ‘Communicable, Maternal, Neonatal, and Nutritional Diseases’ (48,853.6), while our YLD estimates are comparable in magnitude to Non-Hodgkin Lymphoma (682.0) and Idiopathic Developmental Intellectual Disability (642.5) estimates.

### Methodological challenges

There were several methodological challenges observed in this study. Publicly available data was identified through the CSO and the HPSC; however, no data was publicly available collated by sex and age, nor was it available compiled specifically for this study’s date range. To overcome these issues, some necessary assumptions were made. Where data were recorded as “Unknown” (i.e., sex), the ratio from the defined data was applied to the “unknown” data. Age of death was taken as the midpoint in each age category except for the 80+ category where the average age of death was assumed as the average age of persons in that category as per Irish life tables. Privacy restrictions mean that publicly available mortality data with sub-population counts of <5 (i.e., *males 0-14*) remain unavailable, therefore this study’s YLL estimations are applicable only for those aged 24 and above; this in turn will result in DALY underestimation. We estimated a maximum underestimation of 1,118.0 (95%UI 1,113.1, 1,123.2) DALYs. To allow for mortality reclassification, the Department of Health daily briefings were reviewed up to and including 31 March 2021; however, there may have been further reclassification post this date. Despite the above uncertainties, our estimates of COVID-19 mortality are unlikely to significantly differ from the final mortality estimates. Due to the recency and ongoing nature of the COVID-19 pandemic, no attempt was made to account for multimorbidity.

BoD studies use DALYs as a tool to assist international comparability; however, in respect of COVID-19 direct comparability between countries remains difficult as studies may use dissimilar life valuation tables. Additionally, due to existing pre-pandemic vulnerabilities specific to each country’s population, the exact same response applied across countries would likely yield differences in DALY estimates. Therefore, comparisons will not be straightforward, as to achieve the same results proportionately, and differences in investment in interventions would be required.

Comparison of COVID-19 cases with 2019 GBD Study is for contextualisation only. Given the age profile, it is implausible that all COVID-19 deaths are additional (i.e., deaths in the higher age-groups may have occurred irrespectively due to other causes). A lack of clarity remains in relation to ‘dying from COVID’ and ‘dying with COVID’. However, case definitions in RoI have been routinely updated during the pandemic in accordance with ECDC guidance and updates. (8)

This study estimates the direct burden relating to COVID-19, and no attempt was made to measure the indirect burden resulting from COVID-19, or the behavioural changes and implemented control measures (i.e., mental health, reduced screening capacity, missed developmental checks etc).

### Implications for policy and research

COVID-19 has had a detrimental effect on the health of the Irish population. (27, 5) Despite extensive control measures, COVID-19 is likely to be the second highest cause of death from a single disease in the RoI within this study’s duration. It would seem highly likely that the synergistic effects of public health measures implemented during this time period could have had a significant positive population health impact; older adults bore an unequal health burden which ultimately resulted in greater DALYs for this population which were overwhelming informed by YLL. The most obvious strategy for DALY reduction relating to COVID-19 would be to focus on mortality reduction, with particular focus on high-risk groups.

Future research must focus on estimating COVID-19 specific DWs, an extensive BoD study relating to the *indirect* effects of the COVID-19 pandemic, and an extensive study relating to the profile and timeline of “*Long-COVID*”.

## Conclusion

Similar to comparable nations, the health of the Irish population has been impacted by the COVID-19 pandemic, the older populations bearing an unequal burden, particularly in relation to mortality. This work sets out to develop and adapt a standardised COVID-19 BoD methodology framework for Ireland and other comparable EU nations and beyond.

## Data Availability

All data produced in the present study are available upon reasonable request to the authors

## Acknowledgements

This study was completed in association with the European Burden of Disease Network (burden-EU) which was established in 2019 to function as a technical platform for integrating and strengthening capacity in BoD assessment across Europe and beyond. (6)

## Abbreviations

BoD: Burden of Disease
CIDR: Computerised Infectious Disease Reporting
CSO: Central Statistics Office
DALY: Disability Adjusted Life Year
DoH: Department of Health
DPER: Department of Public Expenditure and Reform
DW: Disability Weight
ECDC: European Centre for Disease Control
GBD: Global Burden of Disease
HPSC: Health Protection Surveillance Centre
HSE: Health Service Executive
ICU: Intensive Care Unit
IHME: Institute for Health Metrics and Evaluation
NPHET: National Public Health Emergency Team
PAC: Post-Acute Consequences
PCR: Polymerase Chain Reaction
RLE: Remaining Life Expectancy
RMF: Research Microdata Files
RoI: Republic of Ireland
UCC: University College Cork
UI: Uncertainty Intervals
YLD: Years Lived with Disability
YLL: Years of Life Lost
WHO: World Health Organisation

